# SARS-CoV-2 genomic diversity in households highlights the challenges of sequence-based transmission inference

**DOI:** 10.1101/2022.08.09.22278452

**Authors:** Emily Bendall, Gabriela Paz-Bailey, Gilberto A. Santiago, Christina A. Porucznik, Joseph B. Stanford, Melissa S. Stockwell, Jazmin Duque, Zuha Jeddy, Vic Veguilla, Chelsea Major, Vanessa Rivera-Amill, Melissa A. Rolfes, Fatimah S. Dawood, Adam S. Lauring

## Abstract

**Background:** The reliability of sequence-based inference of SARS-CoV-2 transmission is not clear. Sequence data from infections among household members can define the expected genomic diversity of a virus along a defined transmission chain.

**Methods:** SARS-CoV-2 cases were identified prospectively among 2,369 participants in 706 households. Specimens with an RT-PCR cycle threshold ≤30 underwent whole genome sequencing. Intrahost single nucleotide variants (iSNV) were identified at ≥5% frequency. Phylogenetic trees were used to evaluate the relationship of household and community sequences.

**Results:** There were 178 SARS-CoV-2 cases in 706 households. Among 147 specimens sequenced, 106 yielded a whole genome consensus with coverage suitable for identifying iSNV. Twenty-six households had sequences from multiple cases within 14 days. Consensus sequences were indistinguishable among cases in 15 households, while 11 had ≥1 consensus that differed by 1-2 mutations. Sequences from households and the community were often interspersed on phylogenetic trees. Identification of iSNV improved inference in 2 of 15 households with indistinguishable consensus sequences and 6 of 11 with distinct ones.

**Conclusions:** In multiple infection households, whole genome consensus sequences differed by 0-1 mutations. Identification of shared iSNV occasionally resolved linkage, but the low genomic diversity of SARS-CoV-2 limits the utility of “sequence-only” transmission inference.

**Summary:** High depth of coverage whole genome sequencing can identify SARS-CoV-2 transmission chains in settings where there is strong epidemiologic linkage but is not reliable as a stand-alone method for transmission inference.

## INTRODUCTION

RNA viruses evolve rapidly and accumulate mutations as outbreaks grow [1]. As a result, the evolutionary relationships among sequenced cases hold important information about the processes that drive epidemics [2]. For example, sequence data can help define transmission chains and outbreaks [3–5], the timing and location of viral introductions into communities [6– 8], and larger patterns of spread [9–12]. Over the course of the COVID-19 pandemic, SARS-CoV-2 sequences have been used to infer transmission linkage in hospitals and other congregate settings [13–18]. Inferring these linkages with high confidence is necessary for subsequent studies of the biology of transmission and effectiveness of mitigation strategies.

To infer transmission, one can ask whether the sequences within a group of close contacts, such as a household, are more similar than sequences in the broader community. This approach depends on both the granularity of the sequence data and the amount of genomic diversity in the underlying community or meta-population. The relatedness of viral sequences identified from potential transmission chains versus community virologic surveillance is compared using phylogenetic trees of whole genome consensus sequences or clustering of transmission-associated sequences [2]. In the setting of insufficient community sampling and/or low genomic diversity, consensus trees can miss true linkages and identify false ones. Greater coverage sequencing can improve resolution by identifying intrahost single nucleotide variants (iSNV) in host-derived viral populations that have yet to achieve consensus levels, or >50% within-host frequency, along a transmission chain [19,20]. While these approaches have proven useful for influenza and other viruses, the reliability of sequence-based inference of SARS-CoV-2 transmission is less clear. For example, we and others have found that participants without known epidemiologic linkage can share indistinguishable consensus sequences and even minority (<50%) iSNV [21–23].

Households are ideal settings for studies of the biology and epidemiology of viral transmission. Documentation of close contact and concurrent symptoms or test positivity provide strong epidemiologic evidence of within-household transmission. Sequence data from infected participants can therefore define the expected genomic diversity of a virus along a transmission chain and inform sequence-based studies in other transmission settings, where epidemiologic linkage may be uncertain. Here, we use whole genome sequencing of SARS-CoV-2 populations from participants in two prospective household studies of COVID-19 that were conducted at three sites. To assess the utility of SARS-CoV-2 sequence data as a tool for inferring transmission, we used phylogenetic analysis of sequences from households with at least two SARS-CoV-2 infection cases to assess the clustering of within-household sequences relative to contemporaneous community sequences. We used iSNV to further resolve transmission linkages in selected households.

## METHODS

### Cohorts

The Coronavirus Household Evaluation and Respiratory Testing (C-HEaRT) study enrolled households in Utah (Salt Lake, Weber, Davis, Box Elder, Cache, Tooele, Wasatch, Summit, Utah, and Iron Counties) and New York City [24] during August 2020 through February 2021 and followed them with surveillance for SARS-CoV-2 infection during September 2020 through August 2021. The Communities Organized for the Prevention of Arboviruses (COPA) was expanded to include investigation of the epidemiology of COVID-19, creating the COCOVID study, and recruited households in Ponce, Puerto Rico. For C-HEaRT, household eligibility criteria included: ≥1 child aged 0-17 years, ≥75% of household members met individual level eligibility (all members if a 2-or 3-person household), one adult member was willing to complete monthly questionnaires, and adult members could communicate in English or Spanish. Individual eligibility criteria included: anticipated residence in the household for ≥3 consecutive months, and willingness to complete study surveys, weekly symptom assessments, and self-collect respiratory specimens. For COCOVID, household members were eligible if they were aged ≥1 year, slept in the house ≥4 nights per week, had no definite plans to move in the next year, and were willing and able to comply with study requirements. For both studies, written informed consent (paper or electronic) was obtained from adults (aged >18 years in C-HEaRT and >20 years in COCOVID). Parents or legal guardians of minor children provided written informed consent on behalf of their children; older children (aged 12–17 years in C-HEaRT and 7–20 years in COCOVID) also provided assent to study participation. The C-HEaRT study protocol was reviewed and approved by the University of Utah Institutional Review Board (IRB) as the single IRB for all collaborators. The COCOVID study protocol was reviewed and approved by the Ponce Medical School Foundation, Inc. IRB.

### Sample Collection and Testing

Participants were asked to self-collect (or parent/guardian-collect for children) mid-turbinate nasal swabs every week, regardless of illness symptoms, and place the swabs in viral transport media. Participants were also contacted by text message or email every week to ascertain if they had COVID-19-like (CLI) or any other illness symptoms; they were asked to self-collect an additional mid-turbinate flocked nasal swab once with onset of CLI symptoms. CLI was defined as 1 or more of the following: fever or feverishness, cough, shortness of breath, sore throat, diarrhea, muscle aches, chills, or change in taste or smell. Respiratory specimens were shipped overnight to a central lab and tested using either the Quidel Lyra SARS-CoV-2 Assay or the ThermoFisher Combo Kit platform. The assays were approved under Emergency Use Authorization for the diagnosis of SARS-CoV-2 infection prior to use in this study. Test-positive infections in the same household that were first detected by RT-PCR within 14 days of each other (including those detected on the same date) were considered epidemiologically linked and likely to have resulted from within-household transmission.

### SARS-CoV-2 Genomic Sequencing

SARS-CoV-2 genomic sequencing was attempted on all specimens with an RT-PCR cycle threshold (Ct) ≤30 on either the nucleocapsid protein 1 or 2 target. SARS-CoV-2 genomes were sequenced as described previously [10]. Briefly, RNA was extracted from midturbinate nasal swab specimens with the MagMax MVPII viral nucleic acid isolation kit on a Kingfisher Flex (Thermofisher) and reverse transcribed with Lunascript (NEB). We amplified SARS-CoV-2 cDNA in two pools using the ARTIC Network v3 primers and protocol. Amplicon pools were combined in equal volumes for a given sample and purified with magnetic beads. Barcoded sequencing libraries were prepared using the NEBNext ARTIC SARS-CoV-2 Library Prep Kit with magnetic bead size selection. Individual barcoded sample libraries were pooled (up to 96) and sequenced on an Illumina MiSeq (v2 chemistry, 2×250 cycles).

Reads were mapped to the Wuhan/Hu-1/2019 reference genome (GenBank MN908947.3) with BWA-MEM [25]. We used iVar 1.2.1 [26] to trim ARTIC amplification primer sequences and to determine consensus sequences using bases with >50% frequency and placing a designated unknown base N at positions covered by fewer than 10 reads. Genomes with 29,000 or more unambiguous bases (> 97% completeness) were used in downstream analysis. We identified iSNV with iVar using the following parameters: sample with a minimum consensus genome length of 29,000 bases; sample with an average genome sequencing coverage depth of greater than 200 reads per position; iSNV frequency of 5–95%; read depth of 400 at iSNV sites with a Phred score of >30; iVar p-value of <0.00001. We masked sites commonly affected by sequencing errors in both consensus sequences and iSNV calls [27].

### Phylogenetic Analysis

Consensus sequences for each household were placed on the global SARS-CoV-2 phylogenetic tree using UShER [28]. The tree versions used were from the week of 14 February 2022 and included over 7.8 million genome sequences from GISAID, GenBank, COG-UK and CNCB. The level of genomic sampling of the state or territory of each study site (Figure 1) was estimated with subsampler [10] using case data and GISAID submission data. Subtrees were initially constructed with 30 samples and then reconstructed with additional samples as needed to visualize all genomes from a household in a single subtree (e.g., when samples existed within large clusters of indistinguishable samples). The JSON files for each master tree and subtree are available in Supplemental Dataset 1 and can be visualized in the auspice viewer at https://auspice.us/. Trees were annotated and edited in FigTree using the subtree.nwk files generated by UShER.

**Fig. 1:**
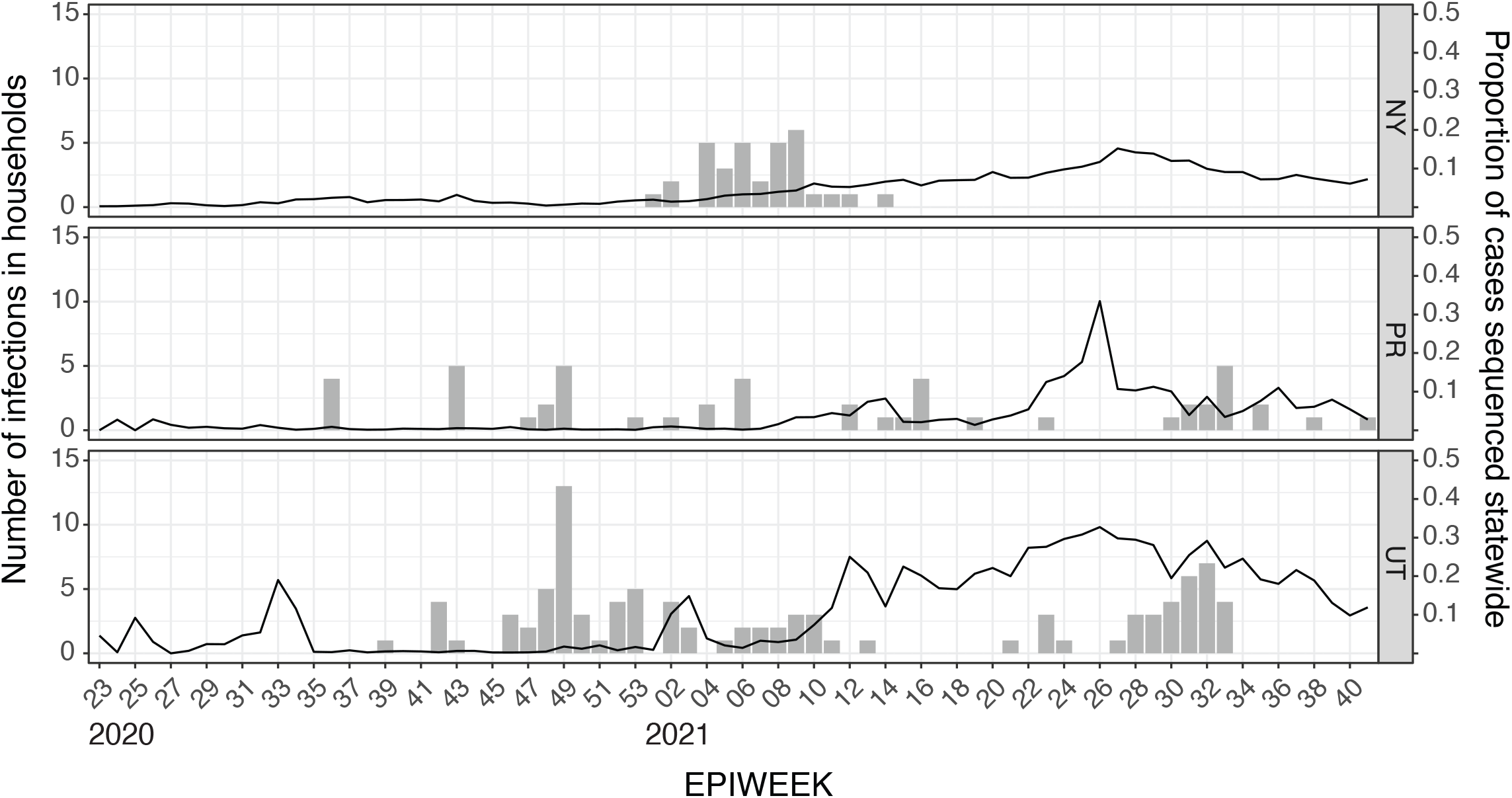
Cases and sampling density. Columns show the number of SARS-CoV-2 infections (left y-axis) in households from New York (NY, top), Puerto Rico (PR, middle), and Utah (UT, bottom) cohorts by epiweek (x-axis). The sampling density (line) for community genomes in each state or territory (right y-axis) was estimated as the proportion of cases with sequences available on GISAID.

### Data and materials availability

The consensus genomes that we generated for this study are publicly available on https://github.com/lauringlab/SARS-CoV-2_Household_Diversity Accessions. Those for the community sequences (largely from GISAID) can be found in the pdf tree files (“<Household_ID>.pdf”) in Supplemental Dataset 1. Laboratories responsible for submissions are acknowledged in Supplemental Table 1. Analysis code for the generation of consensus sequences and phylogenetic analysis are available at https://github.com/lauringlab/SARS-CoV-2_Household_Diversity.

**Table 1:** Distribution of SARS-CoV-2 test-positive cases across cohorts and households

|  | New York<br>City | Utah | Puerto<br>Rico |
| --- | --- | --- | --- |
| Number of Households | 135 | 190 | 381 |
| Number of Participants | 499 | 842 | 1028 |
| Median household size (range) | 4 (2-9) | 4 (2-10) | 2 (1-6) |
| Number of unique SARS-CoV-2-positive cases<sup>a</sup> | 33 | 96 | 49 |
| Households with 1 case | 3 | 17 | 3 |
| Households with 2 cases<sup>b</sup> | 5 | 7 | 5 |
| Households with 3 cases<sup>b</sup> | 3 | 5 | 8 |
| Households with 4 cases<sup>b</sup> | 0 | 4 | 6 |
| Households with 5 cases<sup>b</sup> | 1 | 4 | 5 |
| Households with 6 cases<sup>b</sup> | 1 | 0 | 1 |
| Households with 7 cases<sup>b</sup> | 0 | 1 | 0 |
| # of cases with sequence data / # sequenced | 28/29 | 52/86 | 26/32 |

<sup>a</sup> total number over study period
<sup>b</sup> only households with cases testing positive within 14 days of each other

## RESULTS

The C-HEaRT (Utah and New York City) and COCOVID (Puerto Rico) studies performed active surveillance for SARS-CoV-2 infection and CLI in 706 households with 2,369 participants (Table 1). During September 2020 through August 2021, the cumulative incidence of SARS-CoV-2 infection was 11% [96/842 participants in 41/190 (22%) households under surveillance] at the Utah site and 7% [33/499 participants in 13/135 (10%) households] at the New York City site; during June 2020 through September 2021, cumulative incidence was 5% at the Puerto Rico site (49/1028 infections detected in 28/381 households).

Of the 191 participants with SARS-CoV-2 infections in these households, 147 (77%) in 70 households had samples with a Ct value <30 that were processed for whole genome sequencing, of whom, 106 (72%) had samples that were successfully sequenced to sufficient breadth and depth of coverage (see Methods). Of the 706 households at the three sites, 56 included ≥2 participants who were test-positive within a 14-day period, suggestive of within-household transmission (Table 1). Twenty-six households had high quality sequence data on ≥2 of these contemporaneous infections. The SARS-CoV-2 clades and lineages identified (Table 2) were the same among participants of the same household and reflect viruses circulating in the corresponding time periods in the United States (www.outbreak.info)(Table 2).

**Table 2:** Households with two or more incident SARS-CoV-2 infections within a 14-day period

| Household | Number of Specimens Sequenced | Date First Specimen | Date Last Specimen | Nextclade Clade<sup>a</sup> | PANGO lineage<sup>b</sup> | Mean Consensus Diff<sup>c</sup> |
| --- | --- | --- | --- | --- | --- | --- |
| PR1 | 3 | 9/2/20 | 9/8/20 | 20C | B.1.426 | 0 |
| UT1 | 3 | 10/14/20 | 10/15/20 | 20G | B.1.2 | 0 |
| PR2 | 4 | 10/24/20 | 10/30/20 | 20C | B.1.588 | 0.5 |
| UT2 | 4 | 11/17/20 | 11/24/20 | 20B | B.1.1 | 0.5 |
| UT3 | 6 | 11/24/20 | 12/2/20 | 20G | B.1.2 | 1.4 |
| UT4 | 4 | 11/30/20 | 12/7/20 | 20B | B.1.1 | 1.5 |
| UT5 | 2 | 12/2/20 | 12/3/20 | 20A | B.1.400 | 1 |
| UT6 | 3 | 12/3/20 | 12/10/20 | 20G | B.1.2 | 0.67 |
| PR3 | 4 | 12/3/20 | 12/11/20 | 20B | B.1.1.486 | 1.17 |
| UT7 | 3 | 12/15/20 | 12/28/20 | 20A | B.1.596 | 0 |
| UT8 | 2 | 12/28/20 | 1/1/21 | 21C (Epsilon) | B.1.429 | 0 |
| UT9 | 2 | 1/12/21 | 1/14/21 | 20A | B.1.400 | 0 |
| NY1 | 3 | 1/29/21 | 2/4/21 | 21F (iota) | B.1.526 | 0.67 |
| PR4 | 2 | 2/8/21 | 2/8/21 | 20A | B.1.240 | 0 |
| NY2 | 3 | 2/9/21 | 2/9/21 | 21F (iota) | B.1.526 | 0 |
| NY3 | 3 | 2/11/21 | 2/18/21 | 20C | B.1.582 | 0 |
| NY4 | 2 | 2/21/21 | 3/5/21 | 21F (iota) | B.1.526 | 0 |
| NY5 | 2 | 2/23/21 | 2/23/21 | 21F (iota) | B.1.526 | 0 |
| NY6 | 6 | 2/24/21 | 3/11/21 | 20C | B.1.637 | 1.13 |
| UT10 | 2 | 3/1/21 | 3/15/21 | 21C (Epsilon) | B.1.427 | 2 |
| NY7 | 3 | 3/3/21 | 3/16/21 | 20C | B.1.637 | 0 |
| NY8 | 2 | 3/22/21 | 4/4/21 | 21F (iota) | B.1.526 | 0 |
| PR5 | 2 | 3/23/21 | 3/30/21 | 20B | R.1 | 0 |
| PR6 | 3 | 4/23/21 | 4/23/21 | 20I (Alpha,V1) | Q.4 | 0 |
| UT11 | 3 | 7/26/21 | 8/10/21 | 21J (Delta) | AY.44 | 0 |
| UT12 | 2 | 8/4/21 | 8/4/21 | 21J (Delta) | AY.44 | 1 |

a Defined using nextclade, https://clades.nextstrain.org
b Defined using pango, https://cov-lineages.org/resources/pangolin.html
c Total number of pairwise unambiguous consensus differences between sequences divided by total number of sequences in a household

We first used phylogenetic analysis of whole genome sequences to infer transmission linkage within these 26 households. We used Usher [28] to obtain local sequences for each household and to place household sequences simultaneously on a phylogenetic tree. Over 7.8 million whole genome SARS-CoV-2 sequences were available at the time of this analysis. Because most of these contextual sequences were from GISAID, we estimated the level of sampling at each study site over time by dividing the number of GISAID sequences by the number of reported cases [10]. Sampling of locally circulating viruses was low in 2020 (<2% cases sequenced) and increased at all three sites beginning in early 2021 (>5% cases sequenced, Figure 1). In 2022, Utah was generally better sampled (∼10-30%) than New York or Puerto Rico (5-15%). In 15 out of the 26 households that we studied, the consensus sequences of all cases were indistinguishable and grouped together on their respective trees (representative trees in Figure 2, additional trees in Supplemental Figures 1-4). Given the epidemiologic linkage in the same household, these can be considered sequence-confirmed transmission events. However, we also found several trees in which these monophyletic groupings also included indistinguishable, contemporaneous sequences from non-household members within the community of the same locality (see trees in Figure 2). In two of the New York households, there were many such sequences during a B.1.526 (Iota) variant wave (Supplemental Figures 1 and 2). Therefore, even with a modest level of sampling (Figure 1), it is not uncommon to find indistinguishable viral sequences from participants at the same region and time who presumably lack a documented epidemiologic linkage.

**Fig. 2:**
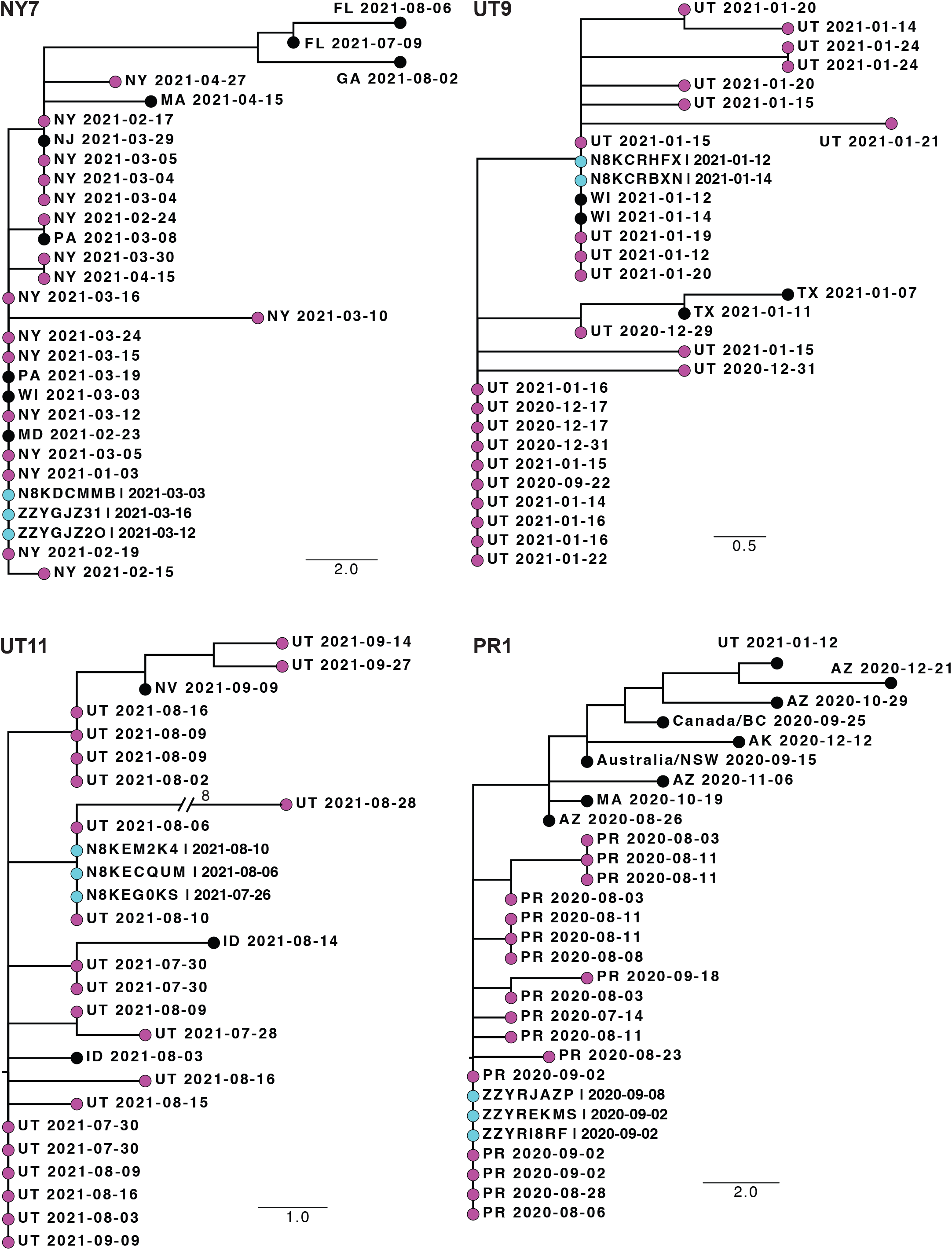
Phylogenetic trees of sequences from households where all participants had indistinguishable consensus sequences. Shown are four representative trees. Trees from 11 other households are shown in Supplemental Figures 1-4. Each tree is labeled with the household identifier (NY = New York, UT = Utah, PR = Puerto Rico). The tips of household sequences are colored cyan and those from non-household participants in the same community in the same state or territory (2 letter abbreviation) are colored magenta. All other tips are colored black. The collection date for each specimen is indicated. Genetic distance is represented by the bar and corresponds to one mutation.

In 11 households, the consensus sequences of the virus from one or more household members differed at 1-2 positions over the nearly 30kb genome. This is not uncommon in transmission chains, particularly ones that are longer or in which the samples are collected 7-14 days apart (see Table 2 for time span). In nearly all cases, the trees from these households demonstrated linkage and/or an ancestor/descendant relationship for the viral sequences (representative trees in Figure 3, additional trees in Supplemental Figure 5). In some cases, the household lineages were phylogenetically distinct from contemporaneous local sequences (e.g., UT2, UT4). These tree structures supported transmission linkage, but low sampling of community cases makes it hard to rule out missed linkages between members of the household and the larger community. Indeed, there were some households in which there were sequences from the larger community included among the same branches of the within-household sequences (e.g., UT10, PR3). As above, the low genomic diversity of SARS-CoV-2 and modest sampling of community cases made it difficult to define a threshold to effectively rule in or rule out transmission.

**Fig. 3:**
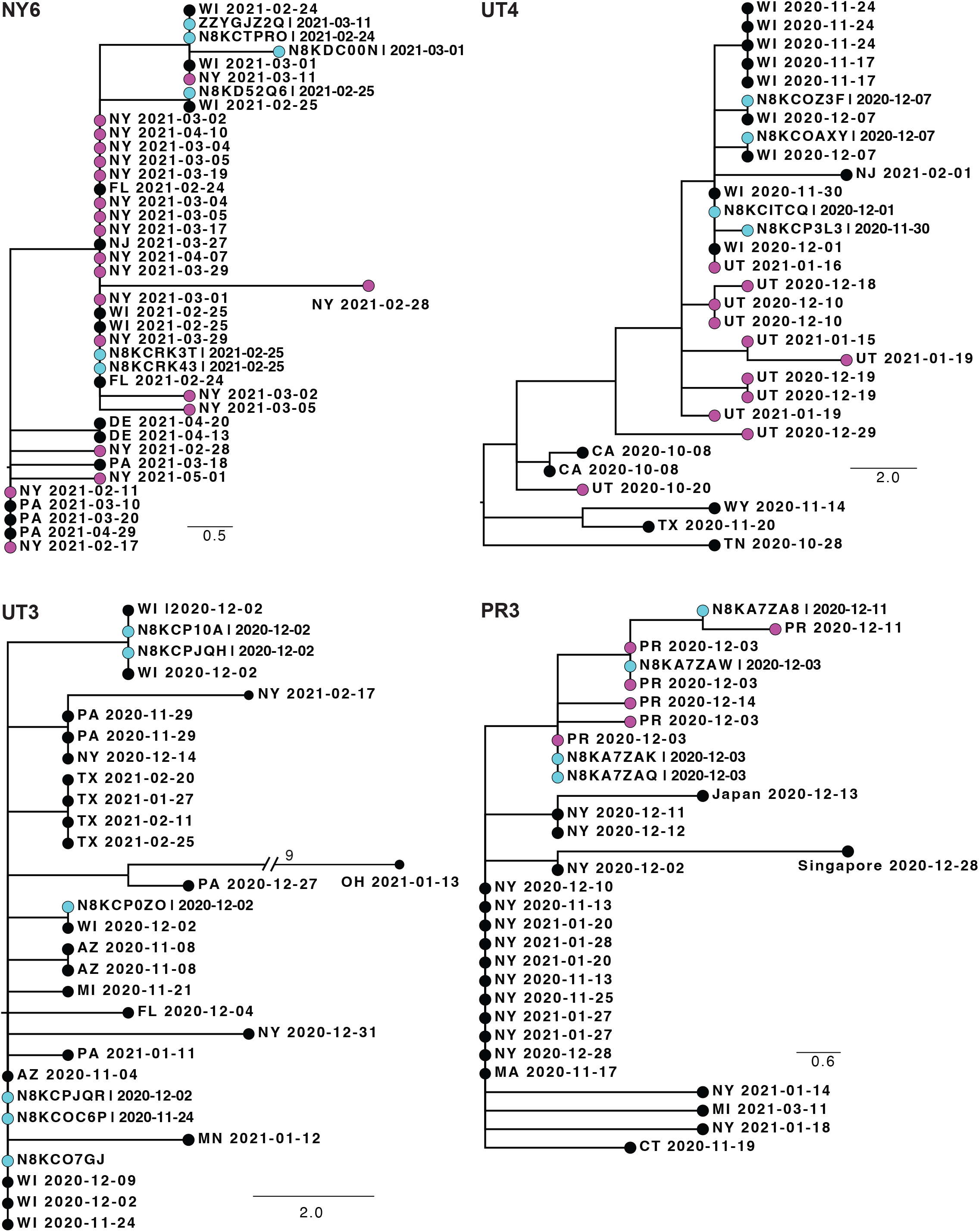
Phylogenetic trees of sequences from households where participants had distinct consensus sequences. Shown are four representative trees. Trees from 7 other households are shown in Supplemental Figure 5. Each tree is labeled with the household identifier (NY = New York, UT = Utah, PR = Puerto Rico). The tips of household sequences are colored cyan and those from the same state or territory (2 letter abbreviation) are colored magenta. All other tips are colored black. The collection date for each sample is indicated. Genetic distance is represented by the bar and corresponds to one mutation.

We next determined whether transmission inference could be improved by identifying iSNV that were shared among members of a household. These would manifest as polymorphic sites where the alternative allele, or mutation, is present but not fixed in the transmission chain. While there was just one household where two participants shared a minority iSNV (PR5, Figure 4), several had iSNV in at least one individual at a consensus level (i.e., frequency >0.5), but that had not yet achieved fixation (i.e., frequency >0.95). In the 15 households with indistinguishable consensus sequences, each of the two participants in households NY5 and PR5 shared an iSNV that was consensus level, but not fixed. In three (UT12, UT5, UT3) out of the 11 households with distinct consensus sequences (Table 2), the consensus differences were due to one or more participants having a non-reference iSNV that achieved consensus level, but not fixation. Household UT4 had two participants with consensus level iSNV (Figure 4). In household PR3, there was one site where one out of four members had a consensus level iSNV and another member had this as a fixed mutation.

**Fig. 4:**
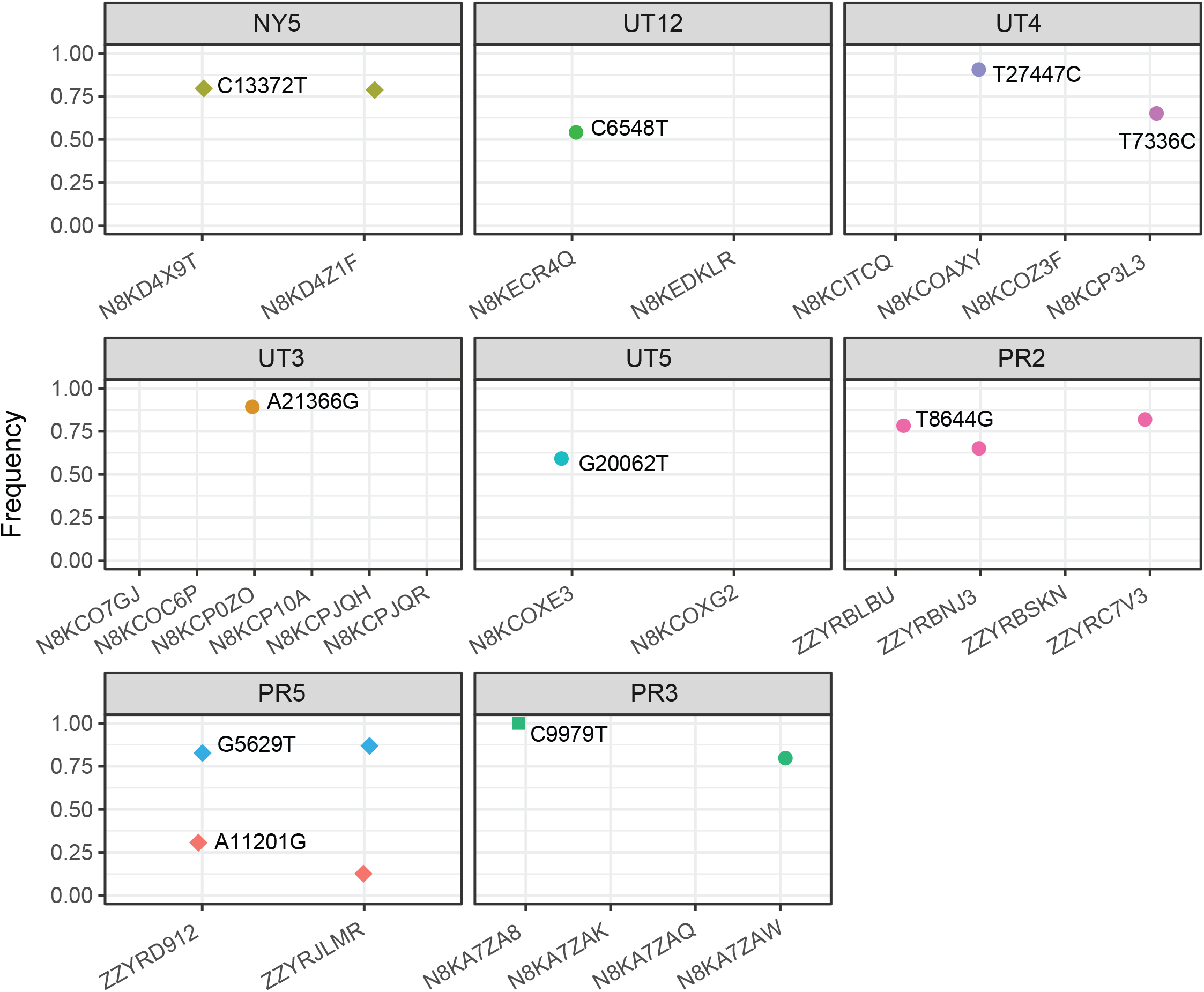
Shared single nucleotide polymorphisms within households. Each panel shows one of the 8 households in which members shared a polymorphic site. The frequency (5-95%) of the indicated mutation (relative to the Wuhan/Hu-1 reference) is shown on the y-axis and the individual/sequence identifier is shown on the x-axis. Shared variants that did and did not lead to a consensus level difference between household members are shown as circles and diamonds, respectively. Mutations that are fixed (>95% frequency) are shown as squares.

## DISCUSSION

We evaluated the utility of SARS-CoV-2 sequence data in transmission inference using data from two studies of household cohorts at three sites. In the household setting, where at least two incident infections occurring within 14 days of one another are strongly suggestive of transmission, we found that sequencing generally confirmed transmission linkage. The whole genome consensus sequences of participants within a household were nearly always indistinguishable or differed by one mutation. In some cases, these links were further supported by the identification of iSNV shared among members of the household. Out of the 26 households evaluated, there was just one (UT10) in which the high average number of consensus differences (two) and absence of shared iSNV called linkage into doubt. Importantly, we frequently found multiple sequences from the community that were indistinguishable to those within the household with even modest sampling (<5%) over the course of the pandemic. This highlights the limits of “sequence-only” inference of transmission in hospitals or other congregate settings where epidemiologic linkage is less certain.

Strengths of the study include our reliance on samples from active surveillance of longitudinal cohorts and our use of quality controlled, deep sequencing. With weekly sampling of all participants from a household, we were able to identify asymptomatic or mildly symptomatic cases and avoid some of the bias of case-ascertainment studies, in which cases are recruited based on a test-positive index. Together with our use of contemporaneous community specimens collected from participants not in the households but from the same site, our data provide a valuable benchmark for the expected SARS-CoV-2 diversity in households relative to that in the community. The cohorts are also drawn from diverse geographic areas with varied household sizes and composition [21,29]. Our assessment of viral diversity is strengthened by our criteria for identifying consensus and minority iSNV [22]. The low observed diversity in this study, in part, reflects the stringent thresholds applied to the sequence data. This conservative approach reduces sequencing errors, which can be systematic and lead to incorrect ascertainment of shared iSNV among unrelated participants [21–23,30].

This study had several notable limitations. First, we were relatively stringent in our criteria for identifying iSNV and therefore may have underascertained shared diversity in the rare (<5%) variant fraction within households. Second, given the limited number of households with sequenced cases, we were unable to formulate a statistically robust approach to sequence-based inference with clear cut-offs and associated positive and negative predictive values. Case-ascertained cohorts or contact tracing studies offer a more efficient way to capture and sequence many putative transmission pairs and will be useful as a setting in which to further develop this approach. Third, while we believe that our data provide an important framework for interpreting sequence data in studies of SARS-CoV-2 transmission, data from households may not translate completely to hospitals and other congregate living settings, which may differ in case density, contact frequency, and force of infection. Fourth, we assumed that household cases testing positive for SARS-CoV-2 within 14 days of one another were linked by transmission. If these cases represented distinct introductions into the household, we could overestimate expected within-household diversity. Fifth, it is possible, but in our opinion unlikely, that some of the community cases in our analysis actually had an epidemiologic linkage to participants in these households.

Despite the limitations identified in this study, integration of sequence and epidemiologic data can be a powerful approach to studies of SARS-CoV-2 transmission. In settings where there is strong epidemiologic linkage among cases (e.g., known exposure or clear temporal and spatial association), indistinguishable consensus sequences with or without shared iSNV should be confirmatory. In these situations, single mutation differences among consensus sequences in a cluster are not uncommon; mutations can fix along a transmission chain, particularly longer ones over a greater timespan. However, if epidemiologic linkage is less certain, sequence identity can only confirm transmission if the metapopulation is highly sampled and genetically diverse. For example, early in the pandemic when circulating SARS-CoV-2 diversity was low, many inpatients and employees in hospitals were found to share indistinguishable consensus sequences, and even iSNV, without any apparent epidemiologic linkage [22,31]. This contrasts with other studies of hospital outbreaks where the combination of contact tracing and sequence data confirmed suspected transmission chains and identified new ones. We expect that future studies of transmission in households, hospitals, and other congregate settings will benefit from Bayesian methods, which can integrate epidemiologic and sequence data for improved inference [32].

## Data Availability

All data are available online at https://github.com/lauringlab/SARS-CoV-2_Household_Diversity.

## ACKNOWLEDGMENTS

We thank the participants in the C-HEaRT and COCOVID cohorts and all GISAID submitting laboratories. We acknowledge Anderson Britto for developing and suggesting the subsampler tool used to generate Figure 1.

## FIGURE LEGENDS

**Supplemental Figures 1-4:**
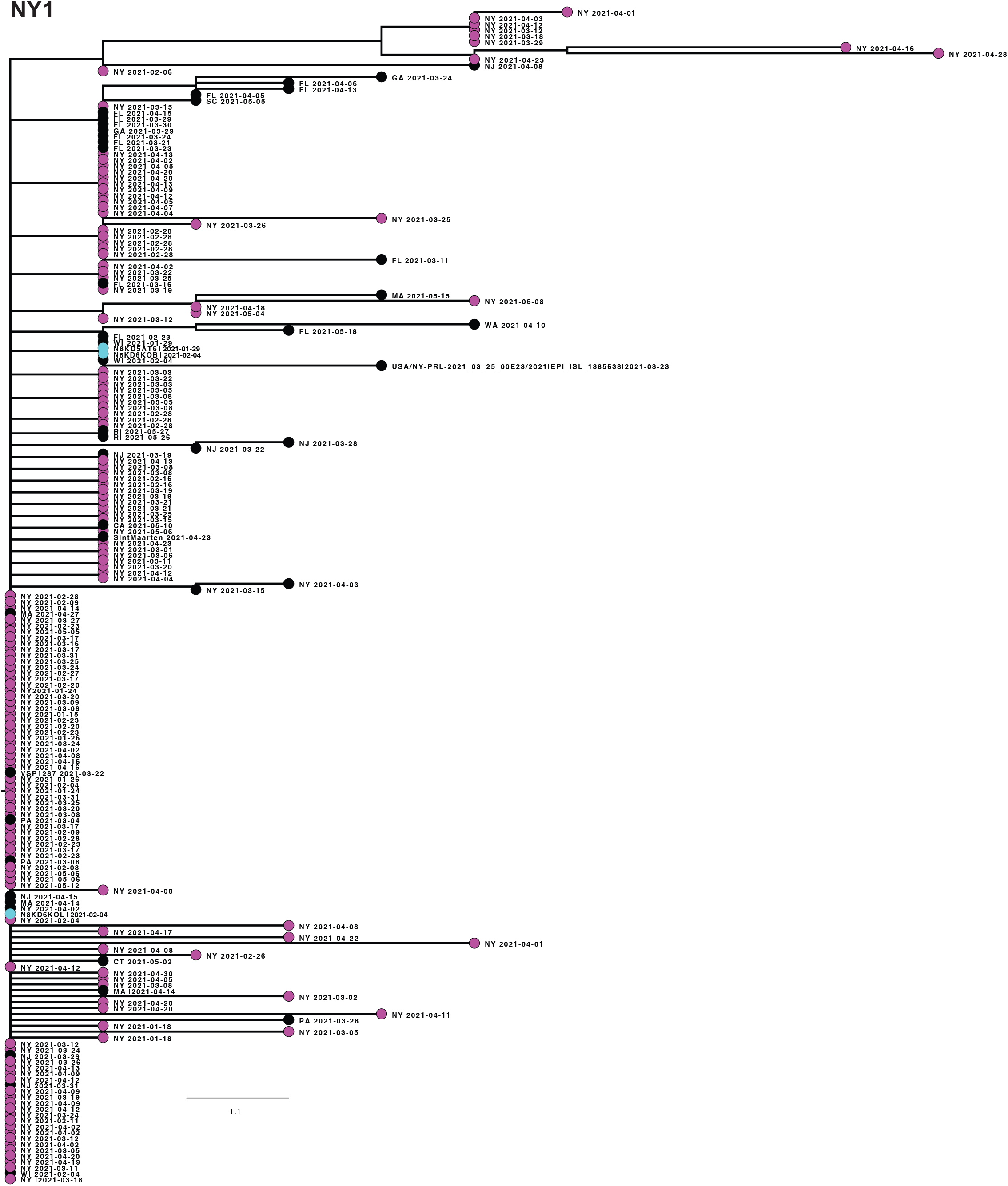
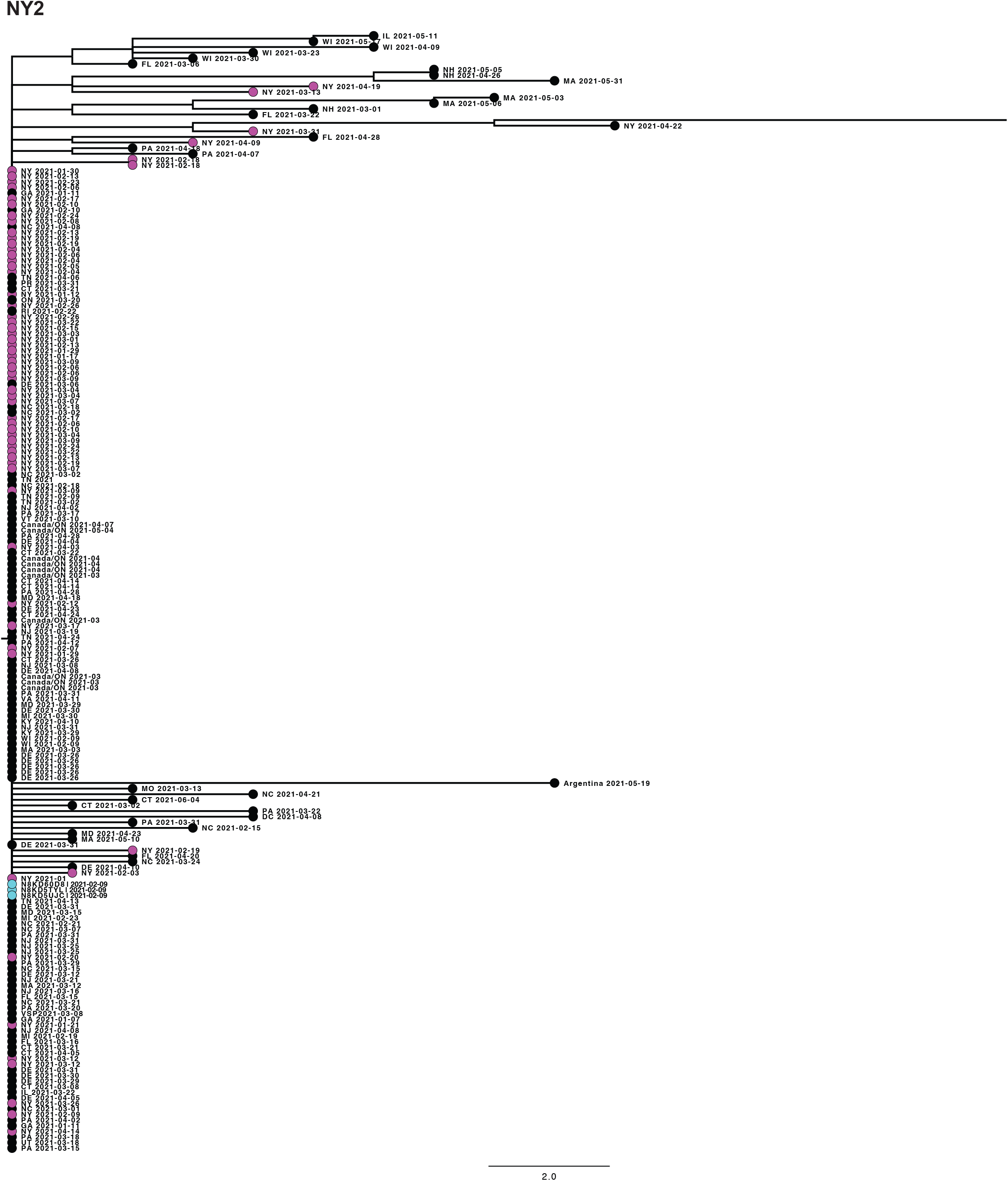
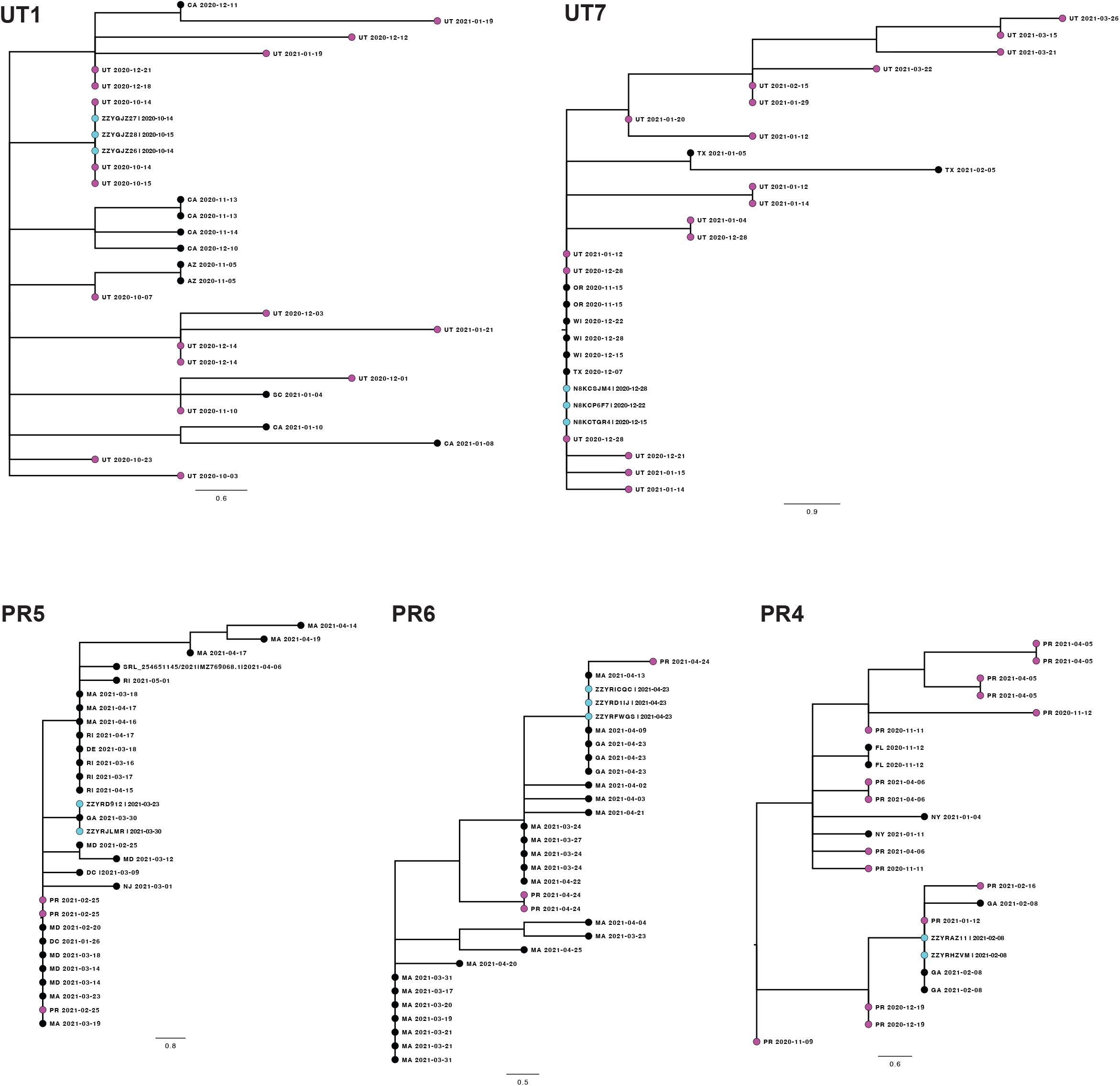
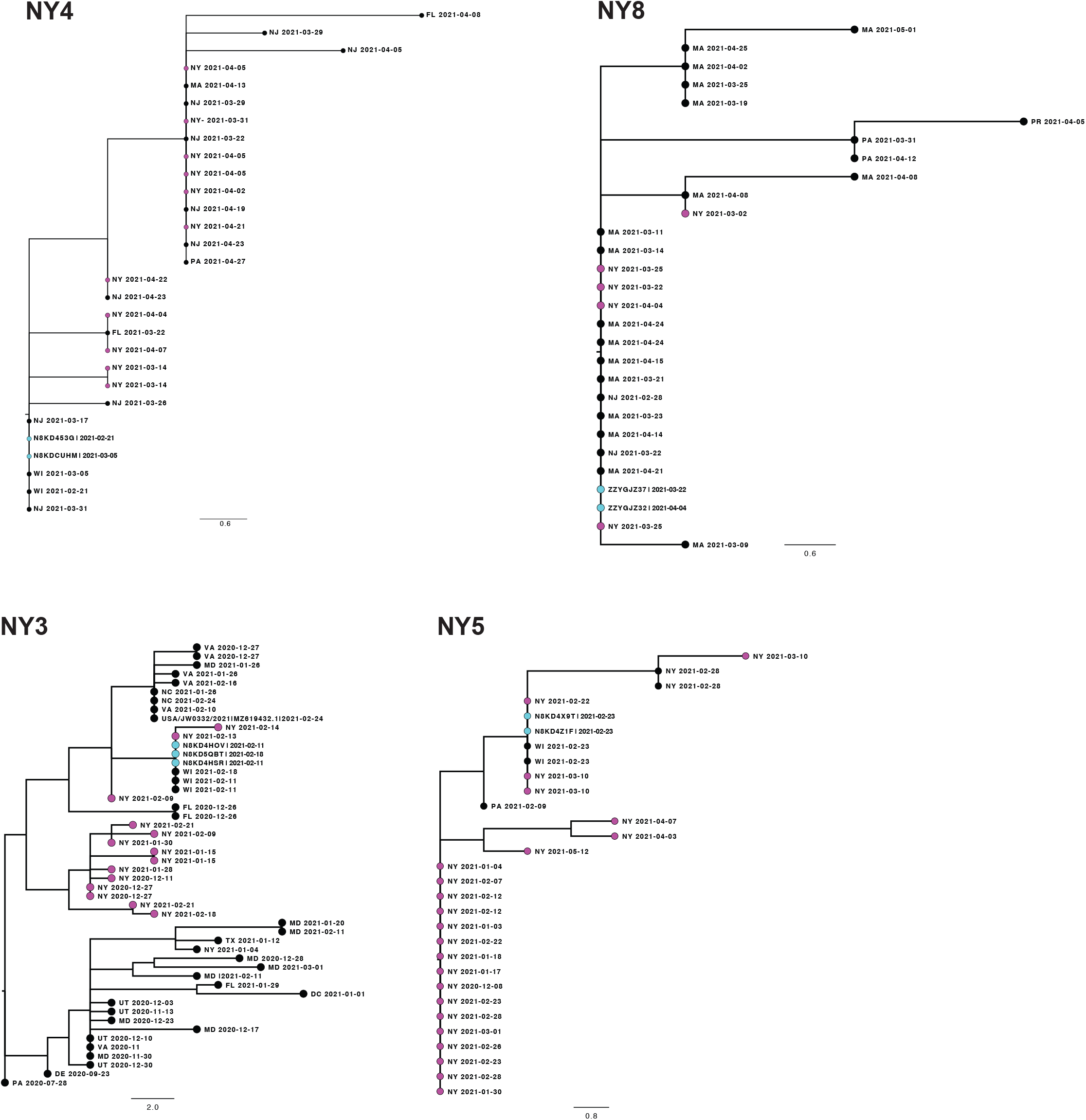
Phylogenetic trees of sequences from households where all participants had indistinguishable consensus sequences. Each tree is labeled with the household identifier (NY = New York, UT = Utah, PR = Puerto Rico). The tips of household sequences are colored cyan and those from the same state or territory (2 letter abbreviation) are colored magenta. All other tips are colored black. The collection date for each sample is indicated. Genetic distance is represented by the bar and corresponds to one mutation.

**Supplemental Figure 5:**
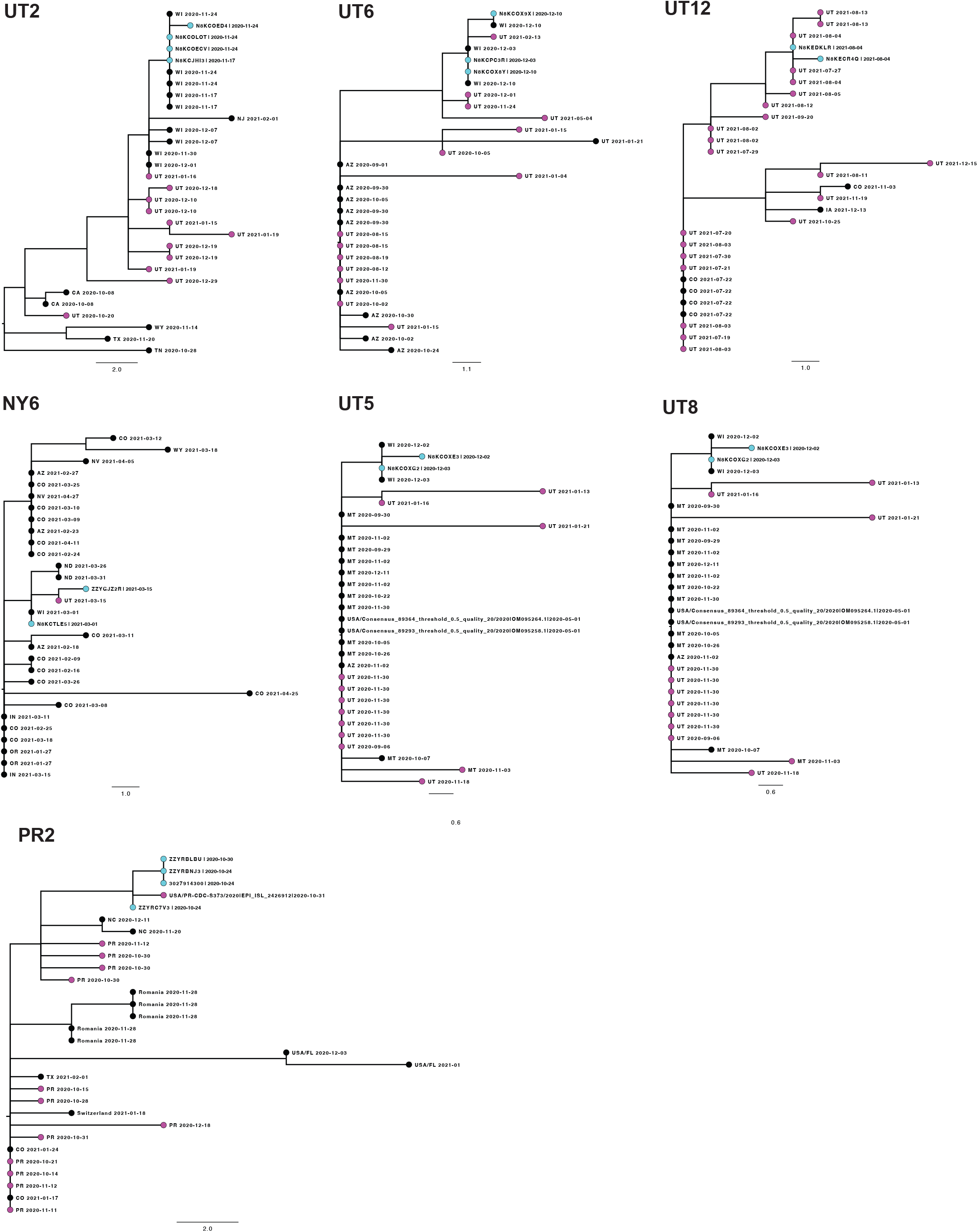
Phylogenetic trees of sequences from seven households where participants had distinct consensus sequences. Each tree is labeled with the household identifier (NY = New York, UT = Utah, PR = Puerto Rico). The tips of household sequences are colored cyan and those from the same state or territory (2 letter abbreviation) are colored magenta. All other tips are colored black. The collection date for each sample is indicated. Genetic distance is represented by the bar and corresponds to one mutation.

**Supplemental Table 1:** Submitting laboratories for GISAID sequences used in this study. GISAID identifiers can be found in the unedited trees in the Supplemental Dataset.

## Notes

### Competing Interest Statement

The authors have declared no competing interest.

### Funding Statement

This work was supported by Centers for Disease Control and Prevention through a contract to Abt Associates Inc.

### Author Declarations

The C-HEaRT study protocol was reviewed and approved by the University of Utah Institutional Review Board (IRB) as the single IRB for all collaborators. The COCOVID study protocol was reviewed and approved by the Ponce Medical School Foundation, Inc. IRB.

### Summary of Updates

Corrected names in the acknowledgements section

